# Risk of Ischemic Stroke in Patients with Covid-19 versus Patients with Influenza

**DOI:** 10.1101/2020.05.18.20105494

**Authors:** Alexander E. Merkler, Neal S. Parikh, Saad Mir, Ajay Gupta, Hooman Kamel, Eaton Lin, Joshua Lantos, Edward J. Schenck, Parag Goyal, Samuel S. Bruce, Joshua Kahan, Kelsey N. Lansdale, Natalie M. LeMoss, Santosh B. Murthy, Philip E. Stieg, Matthew E. Fink, Costantino Iadecola, Alan Z. Segal, Thomas R. Campion, Ivan Diaz, Cenai Zhang, Babak B. Navi

## Abstract

**Key Points:** *Question:* How does the risk of acute ischemic stroke compare between patients with Covid-19 and patients with influenza (a respiratory virus previously linked to stroke)?

*Findings:* In this large retrospective cohort study conducted at two academic hospitals in New York City, patients with emergency department visits and hospitalizations with Covid-19 were approximately seven times as likely to have an acute ischemic stroke as compared to patients with emergency department visits or hospitalizations with influenza.

*Meaning:* Patients with Covid-19 are at heightened risk for acute ischemic stroke as compared to patients with influenza.

*Importance:* Case series without control groups suggest that Covid-19 may cause ischemic stroke, but whether Covid-19 is associated with a higher risk of ischemic stroke than would be expected from a viral respiratory infection is uncertain.

*Objective:* To compare the rate of ischemic stroke between patients with Covid-19 and patients with influenza, a respiratory viral illness previously linked to stroke.

*Design:* A retrospective cohort study.

*Setting:* Two academic hospitals in New York City.

*Participants:* We included adult patients with emergency department visits or hospitalizations with Covid-19 from March 4, 2020 through May 2, 2020. Our comparison cohort included adult patients with emergency department visits or hospitalizations with influenza A or B from January 1, 2016 through May 31, 2018 (calendar years spanning moderate and severe influenza seasons).

*Exposures:* Covid-19 infection confirmed by evidence of severe acute respiratory syndrome coronavirus 2 (SARS-CoV-2) in the nasopharynx by polymerase chain reaction, and laboratory-confirmed influenza A or B.

*Main Outcomes and Measures:* A panel of neurologists adjudicated the primary outcome of acute ischemic stroke and its clinical characteristics, etiological mechanisms, and outcomes. We used logistic regression to compare the proportion of Covid-19 patients with ischemic stroke versus the proportion among patients with influenza.

*Results:* Among 2,132 patients with emergency department visits or hospitalizations with Covid-19, 31 patients (1.5%; 95% confidence interval [CI], 1.0%-2.1%) had an acute ischemic stroke. The median age of patients with stroke was 69 years (interquartile range, 66-78) and 58% were men. Stroke was the reason for hospital presentation in 8 (26%) cases. For our comparison cohort, we identified 1,516 patients with influenza, of whom 0.2% (95% CI, 0.0-0.6%) had an acute ischemic stroke. After adjustment for age, sex, and race, the likelihood of stroke was significantly higher with Covid-19 than with influenza infection (odds ratio, 7.5; 95% CI, 2.3-24.9).

*Conclusions and Relevance:* Approximately 1.5% of patients with emergency department visits or hospitalizations with Covid-19 experienced ischemic stroke, a rate 7.5-fold higher than in patients with influenza. Future studies should investigate the thrombotic mechanisms in Covid-19 in order to determine optimal strategies to prevent disabling complications like ischemic stroke.

## Introduction

Coronavirus Disease 2019 (Covid-19) has affected over 3.5 million people and caused 250,000 deaths worldwide.^1^ Although Covid-19 is primarily a respiratory illness, reports suggest that it may lead to a hypercoagulable state and thrombotic complications.^2-4^ Recent case series from China, France, and New York raise the possibility that Covid-19 might increase the risk of ischemic stroke.^5-7^ However, these studies were small and lacked control groups. To evaluate whether Covid-19 is associated with a higher rate of ischemic stroke than would generally be expected from a viral respiratory infection, we compared the likelihood of acute ischemic stroke in patients with Covid-19 versus patients with influenza, a known stroke risk factor.^8^

## Methods

### Design

We conducted a retrospective cohort study at two hospitals in New York City, one of which is an academic quaternary-care center and the other an academic community hospital. One part of the study population comprised patients aged ≥18 years who had confirmation of severe acute respiratory syndrome coronavirus 2 (SARS-CoV-2) in the nasopharynx by polymerase chain reaction and had an emergency department (ED) visit or hospitalization from March 4, 2020 through May 2, 2020. In parallel, we identified adult patients with an ED visit or hospitalization with laboratory-confirmed influenza A or B at our quaternary-care hospital between January 1, 2016 and May 31, 2018, dates during which we had available data from the Cornell Acute Stroke Academic Registry (CAESAR), which we used to ascertain ischemic strokes in the influenza cohort. Influenza is a common viral respiratory illness that has been established as a risk factor for ischemic stroke,^8,9^ so the comparison between Covid-19 and influenza allowed us to estimate whether Covid-19 is associated with a heightened risk of ischemic stroke beyond that expected from a viral respiratory illness. Calendar years 2016-2018 encompassed both severe (2017-2018) and moderate (2015-2016, 2016-2017) influenza seasons.^10^ Patients with Covid-19 and influenza were identified using automated systems for electronic capture of laboratory results established by the Weill Cornell Medicine Architecture for Research Computing in Health (ARCH) program. Our Institutional Review Board approved this study and waived the requirement for informed consent.

### Measurements

We used automated electronic data capture to collect information on demographics, vascular risk factors, presenting symptoms, severity of Covid-19 illness, laboratory values, imaging studies, medications administered, in-hospital mortality, and discharge disposition.

The primary outcome was acute ischemic stroke. In the Covid-19 cohort, we screened for ischemic stroke by identifying all patients who underwent brain computed tomography (CT) or brain magnetic resonance imaging (MRI) or had an *International Classification of Diseases, 10^th^ Revision, Clinical Modification* (*ICD-10-CM*) diagnosis for cerebrovascular disease (I60-I69) during their ED visit or hospitalization. Two board-certified attending neurologists systematically adjudicated the presence^11^ and etiological mechanism classification^12,13^ of acute ischemic stroke with disagreements resolved by a third independent reviewer. In the influenza cohort, ischemic stroke was ascertained by merging in data from CAESAR. The methods^14^ for stroke adjudication and etiological subtype classification in CAESAR are the same as the methods described above for the Covid-19 cohort.

### Analysis

We used descriptive statistics with exact confidence intervals (CI) to characterize the study population and to calculate proportions of patients with acute ischemic stroke. Comparisons were made using the chi-square test or Wilcoxon rank-sum test for unadjusted comparisons, and logistic regression models were adjusted for age, sex, and race. In a sensitivity analysis, we restricted our Covid-19 and influenza cohorts to patients who were hospitalized. Statistical significance was defined by an alpha-error of 0.05.

## Results

From March 4, 2020 through May 2, 2020, there were 2,132 patients with ED visits or hospitalizations with Covid-19 at our two hospitals. Their median age was 62 years (interquartile range [IQR], 48-75), 55% were men, and vascular comorbidities were common (Table 1).

**Table 1.** Characteristics of Patients with Covid-19, Stratified by the Diagnosis of Acute Ischemic Stroke.

| **Characteristic<sup>a</sup>** | **Acute Ischemic Stroke**<br>**(n=31)** | **No Acute Ischemic Stroke**<br>**(n=2,101)** |
| --- | --- | --- |
| **Demographics** |  |  |
| Age, years | 69 (66-78) | 62 (48-75) |
| Male sex | 18 (58) | 1,159 (55) |
| Race<sup>b</sup> |  |  |
| White | 9 (29) | 602 (29) |
| Black | 3 (10) | 289 (14) |
| Asian | 8 (26) | 268 (13) |
| Other/Unknown | 11 (35) | 942 (45) |
| Hispanic ethnicity | 1 (3) | 397 (19) |
| **Vascular Risk Factors** |  |  |
| Body mass index, kg/m<sup>2</sup> | 27 (24-31) | 28 (23-33) |
| Hypertension | 30 (97) | 1,218 (58) |
| Diabetes | 23 (74) | 843 (40) |
| Hyperlipidemia | 17 (55) | 576 (27) |
| Atrial fibrillation | 17 (55) | 300 (14) |
| Chronic kidney disease | 8 (26) | 323 (15) |
| Coronary artery disease | 16 (52) | 492 (23) |
| COPD | 4 (13) | 190 (9) |
| **Clinical Characteristics** |  |  |
| ICU admission | 18 (58) | 424 (20) |
| Mechanical ventilation | 10 (32) | 296 (14) |
| Prone positioning | 9 (29) | 222 (11) |
| **Laboratory Data** |  |  |
| Initial D-dimer, ng/uL | 1,930 (559-5,285) | 682 (340-1,980) |
| Initial ESR, mm/hr | 89 (60-106) | 71 (45-99) |
| Initial WBC count, 10<sup>3</sup>/uL | 10.3 (6.9-12.9) | 6.9 (5.0-9.6) |
| Initial platelet count, 10<sup>3</sup>/uL | 210 (178-269) | 208 (161-274) |
| Initial troponin-I, ng/mL | 0.03 (0.03-0.09) | 0.03 (0.03-0.06) |

Abbreviations: COPD, chronic obstructive pulmonary disease; ICU, intensive care unit; ESR, erythrocyte sedimentation rate; WBC, white blood cell
<sup>a</sup>Data reported as number (%) for categorical variables and median (interquartile range) for continuous variables.
<sup>b</sup>Percentages were rounded up and therefore many not add to 100%.

Of the 2,132 Covid-19 patients, 31 patients (1.5%; 95% CI, 1.0%-2.1%) had an acute ischemic stroke. The median duration from Covid-19 symptom onset to stroke diagnosis was 16 days (IQR, 5-28) (Figure 1). The median age of patients with acute ischemic stroke was 69 years (IQR, 66-78). Most stroke mechanisms were cardioembolic (n=13, 42%) or cryptogenic (n=16, 52%) (Table 2). Inpatient mortality was 29% among Covid-19 patients with ischemic stroke versus 12% among Covid-19 patients without ischemic stroke (p=0.005).

**Figure 1.**
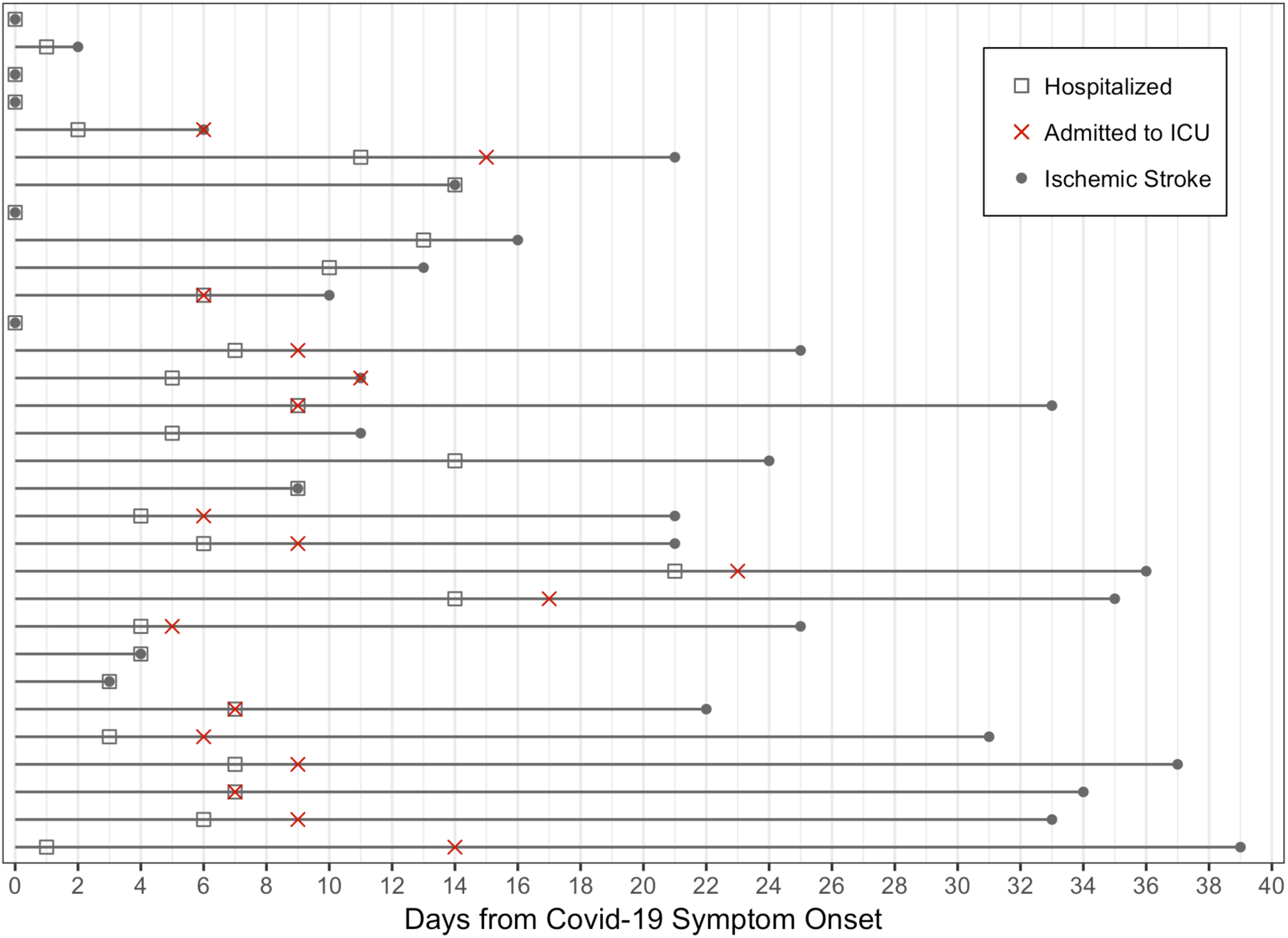
Timeline in days from Covid-19 symptom onset to acute ischemic stroke diagnosis. *Legend:* Horizontal lines represent individual patients with a hospitalization or emergency department visit for Covid-19 infection who had acute ischemic stroke. A gray square indicates the day of hospitalization, a red "X" indicates the day of intensive care unit admission, if applicable, and a gray circle indicates the day of acute ischemic stroke diagnosis. For patients who did not have preceding typical Covid-19 symptoms, the day of their stroke was considered the day of Covid-19 symptom onset. For patients with typical symptoms of Covid-19 but without clear onset date, the date of hospital presentation was considered the day of onset.

**Table 2.**
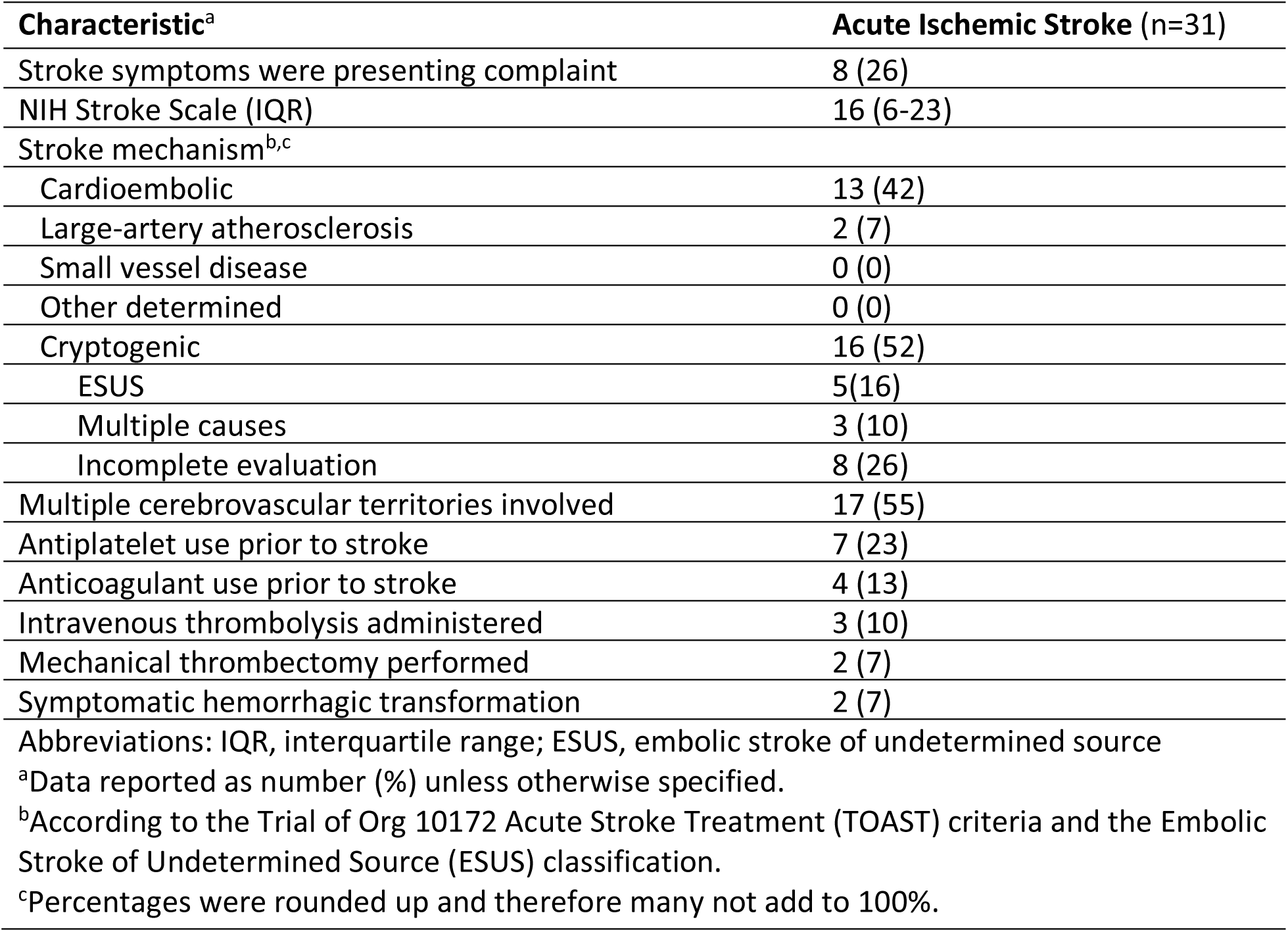
Characteristics of Acute Ischemic Stroke Among Patients with Covid-19.

Among 1,516 patients with influenza from January 1, 2016 through May 31, 2018, three patients (0.2%; 95% CI, 0.0%-0.6%) had an acute ischemic stroke (two cardioembolic and one cryptogenic). In unadjusted analysis, patients with Covid-19 were more likely to have an ischemic stroke than patients with influenza (OR, 7.4; 95% CI, 2.3-24.4). Our results were unchanged after adjustment for age, sex, and race (OR, 7.5; 95% CI, 2.3-24.9). Our results were similar in a sensitivity analysis restricted to hospitalized patients with Covid-19 or influenza (OR, 6.4; 95% CI, 1.9-21.2).

## Discussion

Among 2,132 patients with ED visits or hospitalizations with Covid-19 infection at two major hospitals in New York City, the rate of ischemic stroke was 1.5%, which was 7.5-fold higher than the rate of ischemic stroke among patients who visited the ED or were hospitalized with influenza.

Apart from small case series, our understanding of neurological complications of Covid-19 is limited. Among 214 patients hospitalized with Covid-19 in Wuhan, China, 3% had a stroke.^7^ Among 13 patients with Covid-19 infection who underwent brain MRI in France, 23% had an ischemic stroke.^6^ More recently, investigators reported on five young patients in New York City with cerebral large-vessel occlusion and severe stroke.^5^

Our study of 2,132 racially diverse patients provides a more precise estimate of the absolute risk of ischemic stroke among patients seeking emergency care for Covid-19. Our data indicate that a higher proportion of patients with ED visits or hospitalizations for Covid-19 have acute ischemic stroke compared to patients with ED visits or hospitalizations for influenza.

Further, our findings improve our understanding of the characteristics, mechanisms, and short-term outcomes of ischemic stroke in these patients. We found that most strokes occurred in older age groups and in those with traditional stroke risk factors. We also noted that initial plasma D-dimer levels were nearly three-fold higher in those diagnosed with ischemic stroke.

Our study has several limitations. First, we may have underestimated the true rate of ischemic stroke in patients hospitalized with Covid-19 infection because these patients are sometimes too unstable to undergo brain imaging. In addition, some patients with undiagnosed Covid-19 and stroke may have died before reaching the hospital. Conversely, the recent surge in Covid-19 in New York City could have affected the threshold for visiting the ED or hospitalization and therefore it is possible that patients who sought emergency care with Covid-19 in our cohort were more severely ill than patients who sought emergency care with influenza in past years. The increased risk of stroke in Covid-19 infection may thus reflect greater severity of underlying illness. Second, our study involved two hospitals and thus our results may not be generalizable to other settings. In particular, we were unable to estimate the population-level incidence of ischemic stroke among patients infected with SARS-CoV-2 and compare this incidence to the general population.

In conclusion, the proportion of patients with ED visits and hospitalizations with Covid-19 who had acute ischemic stroke was approximately 7.5-fold higher than the proportion seen in patients who visited the ED or were hospitalized with influenza.

## Data Availability

The data that support the findings of this study are available from the corresponding author upon reasonable request.

## Acknowledgments

None.

## Author Contributions

Babak Navi had full access to all the data in the study and takes responsibility for the integrity of the data and the accuracy of the data analysis.

***Study concept and design:*** Merkler, Parikh, Gupta, Kamel, Navi.

***Acquisition of data:*** Merkler, Parikh, Mir, Lin, Lantos, Lansdale, LeMoss, Navi.

***Analysis and interpretation of data:*** Merkler, Parikh, Mir, Gupta, Kamel, Diaz, Zhang, Navi.

***Drafting of the manuscript:*** Merkler, Parikh, Navi.

***Critical revision of the manuscript for important intellectual content:*** Merkler, Parikh, Mir, Gupta, Kamel, Lin, Lantos, Schenck, Goyal, Bruce, Kahan, Lansdale, LeMoss, Murthy, Stieg, Fink, Iadecola, Segal, Campion Jr, Diaz, Zhang, Navi.

***Statistical analysis:*** Zhang.

***Obtained funding:*** Gupta, Kamel, Navi.

***Administrative, technical, or material support:*** Gupta, Kamel, Fink, Iadecola, Navi.

***Study supervision:*** Navi.

## References

1. World Health Organization. Coronavirus (COVID-2019) situaton reports. 2020. https://www.who.int/emergencies/diseases/novel-coronavirus-2019/situation-reports/. Accessed May 1, 2020.

2. Zhang Y, Xiao M, Zhang S, et al. Coagulopathy and Antiphospholipid Antibodies in Patients with Covid-19. NEJM. 2020;382:e38.

3. Klok FA, Kruip M, van der Meer NJM, et al. Incidence of thrombotic complications in critically ill ICU patients with COVID-19 [published online April 10, 2020]. Thromb Res. doi.org/10.1016/j.thromres.2020.04.013

4. Helms J, Tacquard C, Severac F, et al. High risk of thrombosis in patients with severe SARS-CoV-2 infection: a multicenter prospective cohort study [published online May 4, 2020]. Intensive Care Med. doi.org/10.1007/s00134-020-06062-x

5. Oxley TJ, Mocco J, Majidi S, et al. Large-Vessel Stroke as a Presenting Feature of Covid-19 in the Young [Published online April 28, 2020]. NEJM. doi: 10.1056/NEJMc2009787

6. Helms J, Kremer S, Merdji H, et al. Neurologic Features in Severe SARS-CoV-2 Infection [Published online April 15, 2020]. NEJM. doi: 10.1056/NEJMc2008597

7. Mao L, Jin H, Wang M, et al. Neurologic Manifestations of Hospitalized Patients With Coronavirus Disease 2019 in Wuhan, China [Published online April 10, 2020]. JAMA Neurol. doi:10.1001/jamaneurol.2020.1127

8. Boehme AK, Luna J, Kulick ER, Kamel H, Elkind MSV. Influenza-like illness as a trigger for ischemic stroke. Ann Clin Transl Neurol. 2018;5(4):456–463.

9. Elkind MS, Carty CL, O’Meara ES, et al. Hospitalization for infection and risk of acute ischemic stroke: the Cardiovascular Health Study. Stroke 2011;42(7):1851–1856.

10. Centers for Disease Control. How CDC Classifies Flu Severity. https://www.cdc.gov/flu/about/classifies-flu-severity.htm. Accessed May 1, 2020.

11. Sacco RL, Kasner SE, Broderick JP, et al. An updated definition of stroke for the 21st century: a statement for healthcare professionals from the American Heart Association/American Stroke Association. Stroke 2013;44(7):2064–2089.

12. Adams HP, Bendixen BH, Kappelle LJ, et al. Classification of subtype of acute ischemic stroke. Definitions for use in a multicenter clinical trial. TOAST. Trial of Org 10172 in Acute Stroke Treatment. Stroke 1993;24(1):35–41.

13. Hart RG, Diener HC, Coutts SB, et al. Embolic strokes of undetermined source: the case for a new clinical construct. Lancet Neurol 2014;13(4):429–438.

14. Kamel H, Navi BB, Merkler AE, et al. Reclassification of Ischemic Stroke Etiological Subtypes on the Basis of High-Risk Nonstenosing Carotid Plaque. Stroke 2020;51(2):504–510.

